# Long COVID and kidney function: A scoping review protocol

**DOI:** 10.1101/2024.02.29.24303559

**Authors:** Marcella M. Frediani, Heitor S. Ribeiro, Geraldo F. Busatto, Carlos Roberto Ribeiro Carvalho, Emmanuel A. Burdmann, the HCFMUSP COVID-19 Study Group

## Abstract

**Introduction:** The coronavirus disease 2019 (COVID-19) pandemic had catastrophic repercussions worldwide, including short- and long-term effects on multiple organs. The term “long COVID” encompasses signs and symptoms associated with COVID that persist for more than two months. However, the effects of long COVID on kidney function remain poorly understood.

**Objective:** This scoping review aims to describe the effects of long COVID on kidney function and/or kidney-related outcomes.

**Methods:** We will follow the Joanna Briggs Institute methodology and Preferred Reporting Items for Systematic Reviews and Meta-Analyses extension for Scoping Reviews. Based on the PCC framework, we will include studies that investigated long COVID survivors (**P**articipants); describing kidney function and/or kidney-related outcomes (**C**oncept); in all settings and designs (**C**ontext). A comprehensive literature search was performed using the MEDLINE, Embase, and LILACS databases without date or language restrictions from inception until August 2023. Websites, books, and guidelines will also be searched. Observational studies with retrospective, prospective, and case-control designs will be considered. Two independent reviewers will screen titles and abstracts and perform the full-text review. Data extraction will be done by the main reviewer and checked by a second one. Data about the diagnosis of COVID, follow-up time for long symptoms, and kidney function assessment and outcomes will be extracted from selected evidence. The quantitative results will be synthesized and presented in tables and figures along with a narrative summary.

**Ethics and dissemination:** There is no requirement for ethical approval for this scoping review. On completion, it will be published in a peer-reviewed academic journal and presented at a conference.

## Introduction

The coronavirus disease 2019 (COVID-19) pandemic, caused by SARS-CoV-2 infection, has had catastrophic repercussions worldwide. In addition to the ongoing challenges related to acute infections, healthcare institutions face the burden of late sequelae in previously infected individuals. The term “long COVID” or “post COVID-19” encompasses signs and symptoms associated with COVID that develop or continue three months after the initial SARS-CoV-2 infection and persist for more than two months ^1,2^, according to the World Health Organization (WHO), although different definitions have been used.

Long COVID can involve pulmonary and extrapulmonary manifestations, including fatigue, sleep disturbances, and neuropsychiatric, kidney, gastrointestinal, and cardiovascular dysfunction ^1^. Postulated risk factors for long COVID include severe acute disease requiring invasive mechanical ventilation, female sex, obesity, socioeconomic deprivation, and pre-existing disease ^3,4^. However, the mechanism of long COVID remains unknown.

Kidney involvement is common in acute SARS-CoV-2 infection. Previous studies have reported that up to 28% of hospitalized patients develop acute kidney injury (AKI), and 9% require kidney replacement therapy ^5^. Possible explanations for AKI development include direct effects on the kidney, such as endothelial damage from viral entry, complement activation, local inflammation with tubular injury, and collapsing glomerulopathy. In addition, indirect mechanisms such as sepsis, systemic inflammation, use of nephrotoxic drugs, and hypercoagulability may also play a role in AKI development ^6^.

In non-COVID-19 individuals, strong associations between AKI and the development or progression of late chronic kidney disease (CKD) have been demonstrated ^7^. However, little is known about the long-term effects of COVID-19 on kidney function, and whether there is an association between the severity of COVID-19 in the acute phase of infection and the occurrence of AKI during hospitalization.

Overall, a better understanding of the post-COVID-19 impact on CKD development or progression will improve the development of healthcare strategies for COVID-19 survivors. A preliminary search of PROSPERO, MEDLINE, the Cochrane Database of Systematic Reviews, and JBI Evidence Synthesis was conducted and no current or in-progress scoping reviews or systematic reviews on the topic were identified. Therefore, this scoping review aims to describe the effects of long COVID on kidney function. We hypothesize that COVID is associated with a progressive decline in kidney function, which may increase the risk of CKD development or progression.

## Review question

- What are the long-term effects of COVID-19 on kidney function?

## Inclusion criteria

Eligibility criteria based on the PCC framework (**P**articipants; **C**oncept; **C**ontext) is shown in **Table 1** and described in detail below.

**Table 1.** PCC (Participants; Concept; Context) framework.

|  | Definition |
| --- | --- |
| **Participants** | Adult survivors of COVID-19 in a long-term (≥ three months) |
| **Concept** | Kidney function and/or outcomes (e.g., development or progression of CKD) |
| **Context** | All settings and designs, including in-hospital and community settings |

COVID-19, coronavirus disease 2019; CKD, chronic kidney disease.

### Participants

In this scoping review, we will consider evidence from studies that investigated male or female adult survivors (≥18 years) of COVID-19. The COVID-19 diagnosis will be based on the clinical history and confirmation of a positive reverse-transcriptase polymerase chain reaction (RT-PCR) on swabs collected from nasopharyngeal and/or oropharyngeal samples, positive serologic tests, and clinical and tomographic findings compatible with COVID-19. Evidence from patients on maintenance dialysis will be excluded.

### Concept

We will consider any evidence of long COVID describing kidney function and/or kidney-related outcomes. Long COVID will be defined according to the WHO as “*the continuation or development of new symptoms three months after the initial SARS-CoV-2 infection, with these symptoms lasting for at least two months with no other explanation*” ^2^.

Kidney function will be considered if the estimated or measured GFR is assessed using serum creatinine or cystatin C, iohexol, or radioisotopes. Moreover, kidney function biomarkers and urine markers, such as serum creatinine, serum cystatin C, albuminuria, and proteinuria will also be considered. The main long-term kidney outcomes to be considered are i) development of CKD according to the Kidney Disease: Improving Global Outcomes (KDIGO) ^8^ definition or by ICD10 on medical records; ii) progression of CKD; and iii) initiation of kidney replacement therapy (any kind) or kidney transplantation.

### Context

No restrictions will be applied to the settings or design. Evidence from all available sources that investigated kidney function and outcomes of long COVID will be considered, including in-hospital and community settings.

### Types of Sources

All types of observational study designs (*e*.*g*., retrospective, prospective, case-control, etc.) will be considered. Gray literature sources, conference abstracts, websites, books, and guidelines from prominent medical societies will also be searched. On the other hand, reviews, commentaries, and letters to the editor will not be included.

## Methods

The proposed scoping review will be conducted in accordance with the Joanna Briggs Institute (JBI) methodology for scoping reviews ^9^ and the Preferred Reporting Items for Systematic Reviews and Meta-Analyses extension for Scoping Reviews (PRISMA-ScR; Appendix 1) ^10^.

### Search Strategy

We designed a search strategy to mainly locate published evidence by a peer-review process. A preliminary search on MEDLINE was undertaken to identify relevant articles on the topic. The text words contained in the titles and abstracts of the relevant articles, and the index terms used to describe the articles were used to develop a full search strategy for MEDLINE, EMBASE, and LILACS databases. The search strategy, including all identified keywords and index terms, was adapted for each database and/or information source (see Appendix 2). Databases were searched from January 2020 to August 2023. An additional search was also done on the LitCovid database^11^. The reference lists of all the included sources of evidence will be screened for additional studies. We will not apply language restrictions. Sources of unpublished studies and gray literature to be searched include Google Scholar and Google up to the fifth search page. **Figure 1** shows the PRISMA flow diagram ^12^ with the selected evidence from each database.

**Figure 1.**
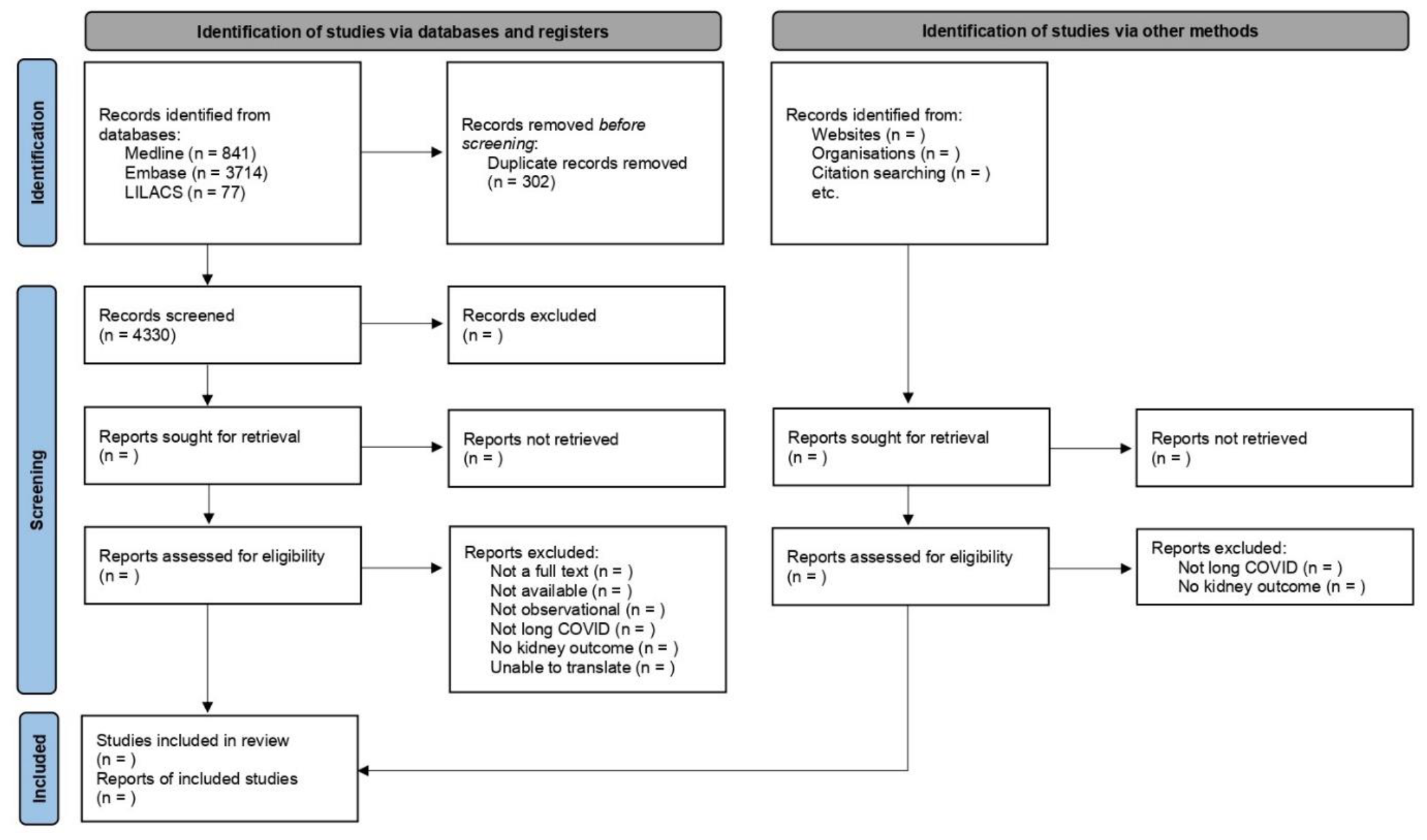
PRISMA 2020 flow diagram

### Source of evidence selection

All identified citations were collated and uploaded into EndNote™ (Clarivate, X9, Philadelphia, PA, USA). Duplicates were removed for the screening process in the Covidence systematic review software (Veritas Health Innovation, Melbourne, AU). Following a pilot test, the titles and abstracts will then be screened by two independent reviewers (MMF and HSR) for assessment against the inclusion criteria. The full text of selected evidence will be assessed in detail by the same independent reviewers. Reasons for the exclusion of sources of evidence in full texts that do not meet the inclusion criteria will be recorded. Any disagreements between the reviewers at each stage of the selection process will be resolved through discussion or with an additional reviewer (EAB). The results of the search and the study inclusion process will be reported in full in the final scoping review and presented in the PRISMA-ScR flow diagram.

### Data extraction

Data will be extracted from the studies included in the scoping review by the main reviewer (MMF) and checked by a second reviewer (HSR) using a data extraction spreadsheet developed by the reviewers, adapted from the JBI Manual for Evidence Synthesis ^13^. The extracted data will include specific details about the diagnosis of COVID, follow-up time for prolonged symptoms, and kidney function assessment. Additional general information will be extracted, including the author(s), year of publication, and country. A draft extraction form is provided (see Appendix 3). The draft data extraction tool will be modified and revised as necessary during the process of extracting data. Any disagreements between the reviewers will be resolved through discussion or with an additional reviewer (EAB). If necessary, the authors of the studies will be contacted to request missing or additional data.

### Quality assessment

Methodological quality assessment of the included studies will be conducted independently by two reviewers (MMF and HSR) using the JBI Critical Appraisal Checklist ^14^. The JBI tool consists of nine items with four possible responses (Yes, No, Unclear, or Not applicable). The greater the number of *yes* answers, the better the quality of the study. We will dichotomize the studies into high-quality (below the median) and low-quality (above the median) for further analysis. Disagreements will be resolved through consensus or with a third reviewer (EAB).

### Data analysis and presentation

The findings from the scoping review will be reported in accordance with the PRISMA-ScR guideline ^10^. Data will be synthesized and presented in tables and figures along with a narrative summary. Statistical analysis will be performed using the Statistical Package for the Social Sciences (version 29.0, IBM Corp., Armonk, NY, USA).

### Patient and public involvement

There will be no patient and public involvement during the review process, however, results will be shared and disseminated throughout extended network within the *Hospital das Clínicas da Universidade de Sao Paulo* (HCFMUSP), which include patients, their relatives, stakeholders, and others.

### Ethics and dissemination

There is no requirement for ethical approval for this scoping review. On completion, it will be published in a peer-reviewed academic journal and presented at a conference.

## Discussion

In this scoping review, we expected to gather as much evidence as possible to comprehensively investigate long-term kidney function in COVID-19 survivors. During the acute phase of infection, the kidney has been demonstrated as a target organ of SARS-CoV-2 ^15^. Despite much evidence in other organs^16,17^, little is known about the impact of COVID on kidney function in the long term. To the best of our knowledge, no previous scoping or systematic review has been addressed this gap. Previous reviews have only highlighted the association between COVID-19 and AKI during hospitalization.

The findings from our scoping review will enlighten kidney function and kidney-related outcomes to be expected in the long-term among those survivors of COVID-19. This may support nephrology healthcare settings on guidance for preventive measures to prevent adverse kidney-related outcomes, such as CKD development and/or progression.

## Supporting information

Appendix 3. Data extraction spreadsheet.

## Data Availability

The datasets to be analyzed during the current study will be available from the corresponding author upon reasonable request.

## Acknowledgments

We would like to thank Editage (www.editage.com) for English language editing.

## Availability of data and materials

The datasets to be analyzed during the current study will be available from the corresponding author on reasonable request.

## Authors’ contributions

MMF, HSR, and EAB conceived and designed the study. MMF and HSR wrote the first draft. GFB, CRRC, and EAB critically reviewed the manuscript. All authors read and approved the final version.

